# Facility-based Pulmonary Rehabilitation Intervention Package (PRIP) for individuals with post-tuberculosis lung disease (PTLD) in Odisha, India: A quasi-experimental study protocol

**DOI:** 10.64898/2026.09.06.26361494

**Authors:** Arohi Chauhan, Abhinav Sinha, Srikanta Kanungo, K Divyasree Bhat, Geetanjali Panda, Shubhangi Shukla, Kirankumar Rade, Sanghamitra Pati

## Abstract

**Background:** India bears the highest global burden of tuberculosis (TB), and despite microbiological cure, a substantial proportion of individuals develop post-tuberculosis lung disease (PTLD), leading to chronic respiratory impairment, reduced functional capacity, and long-term socioeconomic consequences. Although pulmonary rehabilitation (PR) improves exercise capacity and quality of life and is recommended under the National TB Elimination Programme (NTEP), its application remains nascent in routine service delivery, particularly in resource-constrained settings. The key challenge lies not in clinical efficacy, but in the lack of contextually adapted, health system-integrated delivery models. This study aims to evaluate a structured Pulmonary Rehabilitation Intervention Package (PRIP) within public-sector settings in India.

**Methods:** A pre-post quasi-experimental design with a non-randomized comparison group will be conducted over 12 months in two phases. Phase 1 involves adaptation of PRIP through desk review and stakeholder consultations. Phase 2 includes implementation across two districts in Odisha: Khurda (intervention, semi-urban) and Malkangiri (comparator, tribal, resource-limited), minimizing contamination and enabling contextual comparison. Participants with PTLD will receive a 12-week structured PR program including aerobic and strength training, airway clearance, and nutritional counselling. Primary outcome is functional capacity measured by the 6-minute walk test, with secondary outcomes including pulmonary function, dyspnea, symptom burden, quality of life, and healthcare utilization. Implementation outcomes will be assessed using the RE-AIM framework, alongside economic evaluation from health system and societal perspectives.

**Discussion:** This study is designed to evaluate the effectiveness, feasibility, and cost-effectiveness of a structured PRIP for individuals with PTLD within routine public-sector settings in India. Using a quasi-experimental design across two socio-demographically and geographically distinct districts, the study enables a context-sensitive assessment of both intervention outcomes and implementation processes under real-world conditions. By integrating clinical measures, implementation evaluation using the RE-AIM framework, and economic analysis, the study provides comprehensive evidence on scalability and sustainability. The findings will inform integration of PR into the NTEP, particularly in resource-constrained settings. Overall, this study addresses the gap between clinical efficacy and programmatic delivery, supporting a shift from treatment completion to long-term functional recovery and improved quality of life.

**Plain language summary:** Many people who recover from tuberculosis (TB) continue to experience breathing problems, tiredness, and difficulty carrying out everyday activities. This condition, known as post-tuberculosis lung disease (PTLD), can reduce quality of life long after TB treatment has ended. Although pulmonary rehabilitation a program of supervised exercise, breathing techniques, education, and nutritional support has been shown to improve health in people with chronic lung diseases, it is not routinely available for people with PTLD in India.

This study will evaluate a structured Pulmonary Rehabilitation Intervention Package (PRIP) delivered through government health facilities in Odisha, India. People with PTLD will participate in a 12-week rehabilitation program that includes exercise training, breathing exercises, airway clearance techniques, and nutritional counselling. Their progress will be compared with people receiving usual care in another district. The study will assess whether the program improves physical function, breathing, symptoms, quality of life, and healthcare use. It will also examine whether the program is practical, acceptable, and affordable for routine implementation in the public health system.

## Background

Tuberculosis (TB) remains one of the most formidable public health challenges globally, with low- and middle-income countries disproportionately affected.(1) India continues to carry the highest TB burden worldwide, accounting for a substantial share of global incidence and mortality.(2) While advances in diagnostics and standardized treatment regimens have significantly improved cure rates, the long-term health consequences experienced by TB survivors remain unexplored.(3) Increasing evidence indicates that a considerable proportion of individuals who complete TB treatment develop Post-Tuberculosis Lung Disease (PTLD), a chronic condition characterized by persistent respiratory symptoms, impaired pulmonary function, and irreversible structural lung damage.(4) PTLD commonly manifests as chronic cough, dyspnea, reduced exercise tolerance, and functional limitations, leading to sustained morbidity long after microbiological cure.(4)

The impact of PTLD extends beyond clinical outcomes, imposing substantial social and economic burdens on affected individuals and their families.(5) Despite its growing prevalence and impact, PTLD remains underdiagnosed and inadequately managed, reflecting a critical gap in the TB care continuum, which traditionally concludes at treatment completion rather than long-term recovery.(6) Pulmonary rehabilitation (PR) is an evidence-based, multidisciplinary intervention that has demonstrated substantial benefits in chronic respiratory diseases, including improvements in lung function, exercise capacity, symptom control, and health-related quality of life.(7) However, the application of PR for PTLD remains nascent, particularly in resource-constrained settings where structured rehabilitation services are scarce.(7–9) There is a notable lack of context-specific evidence on the feasibility, acceptability, and effectiveness of facility-based PR programs tailored to the needs of PTLD patients.

Given the high burden of TB, the rising prevalence of PTLD, and the absence of structured post-TB rehabilitation services, there is an urgent need to evaluate scalable interventions that extend care beyond microbiological cure. This study seeks to address this critical gap by assessing a structured, facility-based Pulmonary Rehabilitation Intervention Package (PRIP) for individuals with PTLD. By generating robust evidence on its effectiveness and feasibility in high TB-burden settings, the study aims to inform clinical practice and policy, shift the focus from cure to comprehensive recovery, and contribute to sustainable, patient-centered TB care.

## Method

### Study Design

This study will adopt a pre–post quasi-experimental design with a non-randomized comparison group to evaluate the effectiveness and feasibility of a structured PRIP for individuals with PTLD. The study will be conducted over a 12-month period in two phases. Phase 1 will involve adaptation of the PRIP model to the Indian health system through desk reviews and consultations with relevant stakeholders, including clinicians, physiotherapists, and PTLD patients. Phase 2 will involve implementation and evaluation of the intervention in two geographically distinct sites. The intervention arm will be conducted in Khurda district, a semi-urban coastal region with relatively better health infrastructure while the control arm will be carried out in Malkangiri district, a remote tribal-dominated area with limited healthcare access. The ecological and operational separation of these sites will help minimize contamination and allow for a meaningful comparison of outcomes.

### Eligibility criteria

The study population will consist of adolescents and adults aged 14 years and above with a documented history of pulmonary tuberculosis. Eligible participants must have completed anti-TB treatment at least six months before enrolment and present with ongoing respiratory symptoms consistent with PTLD after ruling out active TB. Individuals will be excluded if they have unstable cardiovascular conditions, severe locomotor disability, active malignancy (such as lung cancer), or are moribund or terminally ill. Participants will be recruited from outpatient pulmonary medicine clinics at government healthcare facilities in both districts. Consecutive sampling will be used to enrol eligible patients until the target sample size is achieved.

### Sample size

A sample size of 35 participants per group (intervention and control) has been calculated to detect a mean difference of 55 meters in the 6-minute walk test (6MWT), assuming a standard deviation of 80 meters, with 80% power, a 5% level of significance, and a 10% attrition rate. This brings the total sample size to 70 participants.

### Intervention - PRIP

Participants in the intervention group (Khurda district) will receive a structured pulmonary rehabilitation program over a 12-week period, delivered in twice-weekly supervised sessions. Each session will include aerobic exercises such as free walking and static cycling, strength training exercises like bicep curls, sit-to-stands, pull-ups, and step-ups, airway clearance techniques, and nutritional counseling. Exercise prescriptions will be personalized to maintain oxygen saturation above 85%, monitored through pulse oximetry. Participants will also be encouraged to perform daily home-based exercises using minimal equipment (e.g., water bottles) to support continuity beyond the formal rehabilitation setting. Health education sessions and counseling will be delivered using culturally adapted materials. In contrast, the control group (Malkangiri district) will receive usual care, consisting of routine clinical follow-up and symptomatic management, without structured rehabilitation. After the completion of the study, participants in Malkangiri district will also be offered the PRIP intervention.

### Outcome

Quantitative and qualitative data will be collected at baseline, 6 weeks, and 12 weeks.

#### Primary outcome

The primary outcome is change in walking distance measured by the 6-min walk test from pre-intervention to post-intervention. A group change of at least 54 m is considered clinically important.

6-minute walk distance (6MWD): The 6MWD is a validated measure of functional exercise capacity for chronic respiratory diseases.(10) The 6MWD predicts both future hospitalization and survival.(10) It is responsive to change following pulmonary rehabilitation and is an outcome that matters to patients.(10) The test will be performed in accordance with international standards, including two tests at each time point, with the better (i.e., longest distance) 6MWD recorded.

#### Secondary outcomes

Secondary outcomes will include functional performance using the sit-to-stand test, dyspnea severity using the Medical Research Council (MRC) Dyspnea Scale, symptom burden assessed by the COPD Assessment Test (CAT), quality of life via EQ-5D-5L, healthcare utilization, and cost-benefit analysis.

### Exercise capacity

The five-repetition sit-to-stand test is a commonly used functional performance measure of lower-limb strength.(11) The sit-to-stand test measures the time taken to stand five times from a sitting position as rapidly as possible.(11) It is reliable, valid and responsive to PR with an estimated minimal clinically important difference of 1.7□s.(11)

The MRC dyspnea scale is a 5-point self-administered questionnaire based on the sensation of breathing difficulty experienced by the patient during daily life activities.(12) The questionnaire is short, easy to use and has grades ranging from 1 (none) to 5 (almost compete incapacity), with high grades indicating high perceived respiratory disability.(12) It is responsive to PR with an estimated minimal clinically important difference of 1.(13)

### Health-related quality of Life

EQ-5D-5L: This validated generic quality of life measure is used to estimate health benefits in terms of quality-adjusted life-years (QALYs), and is recommended for economic analyses.(14) It is responsive to PRIP with a minimal clinically important difference of 0.05 (utility index) and 7.0 (Visual Analogue Scale).(15)

### Cost-benefit analysis

The cost of starting and running a PRIP will include single and recurrent costs. Single payments will include the necessary costs needed to set up and run PR. Recurrent costs refer to any item with a life expectancy of ≤1□year (e.g., disposable materials). The fixed costs will be captured prior to enrolling the first participant into the PR programme and the recurrent costs will be collected at the mid-stage of recruitment. The average fixed and recurrent costs will be calculated separately. Additionally, the cost incurred by the participant for PRIP will also be accounted.

Healthcare utilization across 12 weeks: Healthcare utilization will be recorded monthly in participant diaries and telephone record sheets kept by site research staff.

### Data analysis

Data analysis will include descriptive and inferential statistics. Within-group pre-post changes will be analyzed using paired t-tests or Wilcoxon signed-rank tests, and between-group comparisons will use independent t-tests, Mann-Whitney U tests, or ANCOVA to adjust for baseline differences. Repeated measures ANOVA may be used for outcomes measured across all three time points. To evaluate implementation outcomes, the study will apply the Reach, Effectiveness, Adoption, Implementation, and Maintenance (RE-AIM) framework (figure 1).(16) Feasibility, acceptability, and appropriateness will be assessed using validated survey tools. In-depth interviews with patients and healthcare providers will be conducted to explore experiences, barriers, facilitators, and sustainability of the PRIP intervention. Thematic analysis will be used for qualitative data.

**Figure 1.**
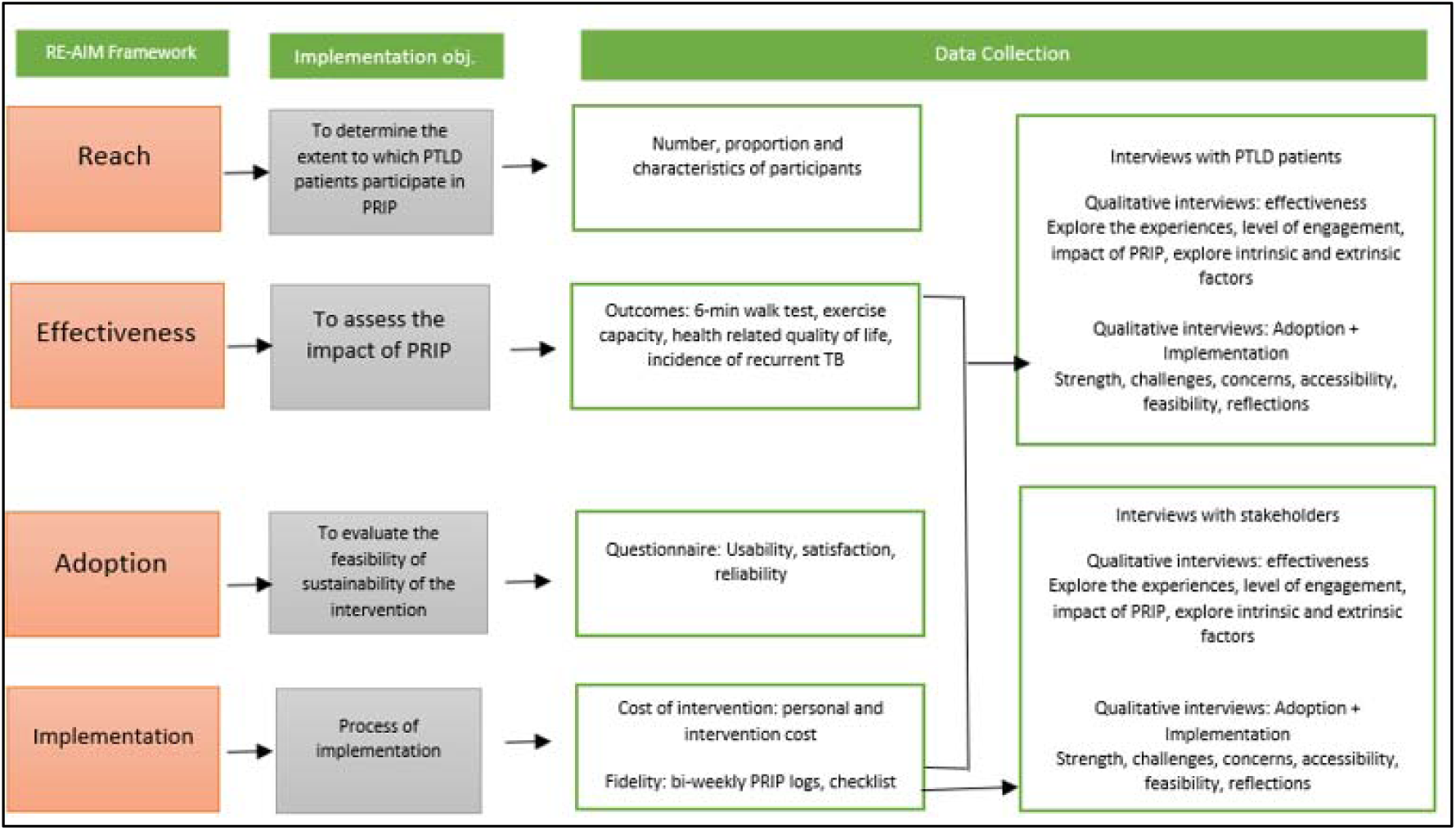
Overview of study objectives with RE-AIM framework.

### Core components of PR exercise

According to the Clinical standards for the assessment, management and rehabilitation of PTLD, the core components of PR program consist of aerobic exercise (endurance training and strength training of upper and lower limbs), airway clearance techniques and nutrition support. The endurance training will include free walking and static cycling. Walking will be individually prescribed at a speed equivalent to >85% SPo2 as measure by pulse oximetry. Walking will be monitored and target duration increased by the instructor as the programme progresses. While the strength exercises will include bicep curls, pull-ups, sit-to-stands and step ups. Individuals will be asked to do one session of strength training and to walk every day at home in addition to structured classes. Minimal equipment will be used where possible, such as bottles filled with water, to achieve the desired weight in kilograms and to offer insight into how participants can exercise effectively at home during and after finishing PRIP. The content delivered within the education sessions will be based on existing materials and nutritional counselling.

### Assessments

It will be done at the baseline while screening for the eligibility criteria as well as after 6-weeks and 12-weeks of the intervention. (Table 1)

**Table 1.** Assessment and follow up.

| Investigation | Baseline | 6-weeks | 12-weeks |
| --- | --- | --- | --- |
| Written informed consent | + |  |  |
| Socio-demographic data | + |  |  |
| Out-patient visits and hospital admissions | + | + | + |
| Medical history | + | + | + |
| Clinical examination | + | + | + |
| Chest x-ray | + |  | + |
| PFT | + | + | + |
| 6-min walk test | + | + | + |
| Medical Research Council Dyspnea scale | + | + | + |
| COPD assessment test | + | + | + |
| EQ-5D-5L | + | + | + |
| Diagnostic test for *M.tb* * | + | + | + |
| Sit to stand test | + | + | + |
| Cost benefit analysis | + | + | + |

\*conducted for those with signs and symptoms suggestive of pulmonary TB

## Discussion

PTLD is emerging as a major long-term sequela in individuals treated for pulmonary TB, yet it remains largely underdiagnosed and undertreated in high-burden countries like India.(17) As TB programs have historically focused on microbiological cure and treatment completion, the chronic respiratory impairments and functional limitations experienced by many TB survivors have not been adequately addressed in routine care.(18) The limited availability of structured PR services, particularly in public sector facilities and rural or tribal areas, exacerbates this care gap.(19) Current evidence supports the benefits of PR in chronic respiratory diseases; however, its role in PTLD has only recently begun to gain attention, and robust evidence from diverse settings within India is lacking.(17)

This study protocol addresses a critical knowledge and implementation gap by evaluating a structured, facility-based PRIP tailored for individuals with PTLD. By employing a quasi-experimental design with a comparison group, the study strengthens causal inference and provides real-world evidence on PRIP effectiveness. Importantly, the study draws on the contrast between two distinct healthcare contexts Khurda (semi-urban) and Malkangiri (tribal) to assess the feasibility, acceptability, and adaptability of PRIP in diverse populations.(20) The integration of both clinical and implementation outcomes, guided by the RE-AIM framework, ensures that the findings will be actionable and relevant to policymakers and program managers.(21)

The use of a comprehensive intervention including aerobic and strength training, airway clearance, and nutrition counselling based on international clinical standards ensures alignment with best practices.(8) Assessing outcomes such as exercise capacity (6MWT), lung function, dyspnea, symptom burden, and quality of life provides a multidimensional understanding of patient recovery.(22, 23) The inclusion of cost-benefit analysis further supports the policy relevance of the study by offering insights into scalability and resource implications.(7)

### Conclusion

This study will generate critical evidence on the clinical effectiveness, feasibility, and implementation potential of a facility-based pulmonary rehabilitation program for PTLD patients in India. By targeting a significant but overlooked component of the TB care continuum, the findings can help shift the paradigm from treatment completion to holistic recovery. If successful, PRIP has the potential to be scaled through district hospitals and health and wellness centres, complementing the National TB Elimination Programme (NTEP) and Universal Health Coverage goals. In the long term, integrating PR into post-TB care pathways could improve health outcomes, reduce disability, and restore productivity in TB survivors especially among vulnerable and underserved populations. This study marks a step toward a more comprehensive, patient-centred approach to TB management in India.

## Declarations

### Ethics approval and consent to participate

Ethical approval has been obtained from the Institutional Ethics Committee of ICMR-Regional Medical Research Centre (RMRC), Bhubaneswar. Written informed consent will be obtained from all participants prior to enrolment. Participant confidentiality will be protected using anonymized data and secure storage systems. Study findings will be disseminated through peer-reviewed publications, policy briefs, and stakeholder consultations, with the aim of informing scalable integration of pulmonary rehabilitation into routine post-TB care services in India.

### Consent for publication

NA

### Availability of data and materials

All the data generated from this study will be available from corresponding author on reasonable request and will also be presented in the manuscript resulting from this study.

### Patient and Public Involvement

Individuals with PTLD will be involved in stakeholder consultations to inform the contextual adaptation and delivery of PRIP. Participant feedback will also inform assessment of its feasibility, acceptability and appropriateness.

### Authors’ contributions

AC, KR and SP conceived and designed the study. AC drafted the study protocol. AS, GP and SS provided administrative and technical support. AS, SK and DB contributed to methodological inputs and critically revised the manuscript for important intellectual content. All authors read and approved the final manuscript.

## Data Availability

All data produced in the present study are available upon reasonable request to the authors

## Acknowledgements

NA

